# Improving parents’ knowledge of early signs of pediatric eye disease: A double-blind RCT

**DOI:** 10.1101/19009183

**Authors:** Sandra E. Staffieri, Gwyneth Rees, Paul G. Sanfilippo, Stephen Cole, David A. Mackey, Alex W. Hewitt

**Affiliations:** Centre for Eye Research Australia, Royal Victorian Eye and Ear Hospital, East Melbourne, VIC, Australia; Ophthalmology, University of Melbourne, Department of Surgery, East Melbourne, VIC, Australia; Royal Women’s Hospital, Parkville, VIC, Australia; Lion’s Eye Institute, Centre for Ophthalmology and Visual Sciences, University of Western Australia, Perth, WA, Australia; Menzies Institute for Medical Research, School of Medicine, University of Tasmania, TAS, Australia

**Author notes:** **Address correspondence to**: Dr Sandra Staffieri. Centre for Eye Research Australia 32 Gisborne Street, East Melbourne 3002 Victoria, AUSTRALIA. **Financial disclosure:** The authors have no financial disclosures to declare. **Clinical Trial Registration:** ANZCTR.org.au identifier: ACTRN12617001431314p; World Health Organization Universal Trial Number: U1111-1203-0485. **Data Sharing Statement:** Deidentified individual participant data (including data dictionaries) will be made available upon request, in addition to study protocols, the statistical analysis plan, and the informed consent form. The data will be made available upon publication to researchers who provide a methodologically sound proposal for use in achieving the goals of the approved proposal. Proposals should be submitted to. **Contributors’ Statement** Dr Staffieri conceptualized and designed the study; developed and designed the intervention content and evaluation surveys; recruited all participants and administered evaluation surveys; collected, analysed and interpreted the data; drafted the initial manuscript and revised the final manuscript. Professor Hewitt, Professor Mackey and Dr Rees critically reviewed the study design, intervention content and evaluation surveys; critically reviewed the manuscript for important intellectual content. Dr Sanfilippo revised the evaluation survey, completed statistical analyses and critically reviewed the manuscript for important intellectual content. Dr Cole critically reviewed the study design and critically reviewed the manuscript for important intellectual content. All authors approved the final manuscript as submitted and agree to be accountable for all aspects of the work.

## Abstract

**Background and Objectives:** Early diagnosis and intervention is essential to achieve optimal outcomes for most pediatric eye diseases. Educating parents/caregivers to recognize early signs of disease and consult a healthcare professional is critical to achieving this aim. We evaluate the effectiveness of an eye-health information pamphlet on parents’ level of concern and their help-seeking intention if they observed leukocoria or strabismus.

**Methods:** Pregnant women attending a metropolitan antenatal clinic were recruited to the study. Participants were randomly assigned to receive a pamphlet on either pediatric eye health (intervention) or strategies for play (control). The primary outcome measure was a change in the parents’ level of concern if they observed leukocoria or strabismus. The secondary outcome measure was a change in their help-seeking intention if either sign was observed.

**Results:** Of the 518 women enrolled, 382 (73.7%) completed the post-test survey. At follow-up, women who received the intervention were more likely to report a higher level of concern if they observed leukocoria (OR 1.711 [CI: 1.176-2.497] p=0.005]) and were less likely to have a delayed help-seeking intention. (OR 0.560 [CI 0.382-0.817] p =0.003) No change in the level of concern for strabismus was identified between the groups; however, at follow-up, women who received the intervention were less likely to delay help-seeking (OR 0.318 [CI 0.125-0.806] p=0.016).

**Conclusion:** Providing parents with relevant, evidence-based information can significantly improve their knowledge and positively influence help-seeking intentions if leukocoria or strabismus are observed.

**Trial registration:** ANZCTR.org.au identifier: ACTRN12617001431314p;

World Health Organization Universal Trial Number: U1111-1203-0485

**Table of Contents Summary:** This study reports the results of a randomised controlled trial evaluating a novel, evidence-based, theory-informed pediatric eye-health information pamphlet for parents.

**What is known on this subject:** Lack of parental awareness of signs of pediatric eye disease (leukocoria and strabismus) delays consultation with healthcare professionals (help-seeking), contributing to late diagnosis and poor outcomes. Providing parents with relevant health information can improve their child’s health outcomes.

**What this study adds:** Using an RCT to evaluate a novel health intervention, this study demonstrates that providing parents with evidence-based, theory informed pediatric eye-health information can improve their knowledge and help-seeking intentions if leukocoria or strabismus are observed in their child.

## BACKGROUND

Pediatric eye disease can range from relatively minor (e.g. congenital nasolacrimal duct obstruction) to potentially blinding (e.g. cataract) or fatal. (e.g. retinoblastoma). Generally, parents (or primary caregivers) will be the first to observe any direct clinical sign or symptom associated with these diseases prompting consultation with a health-care professional (help-seeking). To ensure timely diagnosis and treatment of pediatric eye disease, parents must be aware otherwise asymptomatic signs that can occur during their child’s early infancy and childhood, and where and when to seek eye-health consultation.

Systemically asymptomatic signs such as leukocoria (white pupil) and strabismus (crossed/turned eye) are commonly associated with significant eye disease such as cataract and retinoblastoma.^1, 2^ Leukocoria can easily be dismissed by parents due to the absence of concomitant physical symptoms; when it is only observed intermittently; or is only apparent in photographs (photo-leukocoria) and thought to arise as a photographic artefact.^3^

Although strabismus is usually idiopathic with a prevalence of between 1-6%^4–7^, undiagnosed and untreated it leads to amblyopia and abnormal development of stereopsis.^4, 5^ Less frequently, but more significantly, strabismus develops secondary to severe vision loss^8^; systemic disease^9^; or to intracranial^10^ or intraocular pathology.^1^ Whilst intermittent or even constant strabismus is a common finding in healthy newborns, normal binocular co-ordination should be achieved by approximately 4 months of age.^11, 12^ Any persisting strabismus after this time should be investigated to exclude any serious secondary disease.

Currently, parents in Victoria, Australia are not provided with any information regarding signs of paediatric eye disease to be alert to. There is a pressing need to provide relevant information regarding ocular development during childhood and signs not to be ignored. The aim of this study was to evaluate an eye-health information pamphlet specifically designed to improve knowledge and help-seeking intention if parents observed strabismus or leukocoria in their children.

## METHODS

### Study design

Using a pre- /post-test design, this double-blind, randomized controlled trial (RCT) compared participants’ responses to two clinical scenarios and statements of belief when faced with observing leukocoria or strabismus in a child. This study was approved by the Royal Women’s Hospital Human Research Ethics Committee (#17-38) and adhered to the tenets of the Declaration of Helsinki. Following study commencement, no significant changes were made to the study protocol.

### Participant eligibility criteria and study setting

Participants were recruited from the antenatal outpatient clinic in a large metropolitan hospital in Victoria, Australia. Prima- or multiparous women aged over 18 years in their 2nd or 3rd trimester of pregnancy were invited to participate in the study. The study materials were only available in English.

### Intervention and Control Arms

The intervention consisted of a 4-sided, A5 information pamphlet for parents describing normal developmental milestones for infant vision and ocular development; information about strabismus and leukocoria; and recommendations for seeking health advice if observed. The pamphlet as described in Supplementary Materials A, was specifically designed using an evidence-based approach to developing health promotion materials^14^ and grounded in a theoretical model for behaviour change.^13, 14^ The control group received an A4 double-sided information pamphlet for parents: “Playing with your baby”. This pamphlet encourages strategies for engagement with a newborn and is described in Supplementary Materials B.

### Adherence to the intervention

Following informed consent, completion of the baseline survey and randomization (0-time), participants in both groups were verbally advised by the recruiting researcher SES to read the instructions and pamphlet enclosed in their allocated, sealed research envelope. Two-weeks after baseline (0-time + 2 weeks), participants were sent a secure link by email to complete the follow-up survey using the Research Electronic Data Capture tool (REDCap 7.2.2 © Vanderbilt University). Reminder emails were sent at 0-time + 4 weeks, and 0-time + 5 weeks and a final telephone call was made at 0-time + 6 weeks. Non-responders at the close of the study were recorded as being lost to follow-up. No inducements were offered although at enrolment, participants were advised that those who at the conclusion of the trial were found to have been randomized to the control group, would also receive the intervention.

### Measures and Outcomes

At baseline, participants answered socio-demographic questions pertaining to age, previous experience with infant development, primary language spoken at home, educational attainment and professional background. Participant health literacy was measured by embedding the five questions of Domain 9 “Understanding Health Information” of the validated Health Literacy Questionnaire (HLQ).^15^

The evaluation survey comprised four- or five-point Likert scale items and single item True/False questions specifically developed for six clinical scenarios and 20 statements of knowledge or belief. To maintain masking of the participants to the specific focus of the study and aiming to minimize any additional search for information they would not otherwise be prompted to undertake after baseline not all questions related to infant vision or ocular health.

To evaluate participants’ knowledge and help-seeking intentions if they observed leukocoria or strabismus in their child, a single clinical scenario and statement of belief each pertaining to leukocoria and strabismus were embedded in the survey. Socio-demographic questions were excluded from the follow-up survey and replaced with questions examining participants’ pamphlet engagement. To account for any other sources of information that may contribute to their responses in the follow-up survey, questions relating to searching for more information were also included. (Supplementary materials C) Either baseline or follow-up surveys took less than 10 minutes to complete.

### Outcomes

The primary outcome measure for this study was a change in the level of concern if the participant observed leukocoria or strabismus as measured on a 4-point Likert scale for a clinical scenario and a statement of belief. For example: *“How concerned are you?”* a) not at all concerned; b) somewhat concerned; c) concerned; d) very concerned; and in response to the statements: *“I would be concerned if I saw a white pupil in a photograph of my child”* or *“I would be concerned if noticed my 2-year-old baby’s eye turning (cross-eyed) some of the time”*: a) strongly agree; b) agree; c) disagree; d) strongly disagree. The secondary outcome measure was a change in help-seeking intention if the parent observed either leukocoria or strabismus as measured on a 5-point Likert scale. *“How quickly would you seek advice?”* a) today/tomorrow; b) within 1 week; c) within 1 month; d) longer than 1 month; e) not at all. We hypothesized that women receiving the intervention would demonstrate an increase in concern if leukocoria or strabismus were observed and demonstrate a change in their help-seeking intention.

### Sample size and power calculation

Using STATA (Release 14, 2015),^16^ a sample size of 398 participants (199 in each group) was calculated to confer 80% power to demonstrate a 10% effect with a two-sided p=0.05 as we could not be certain of the direction of the intervention effect. Allowing for a 30% attrition rate, the study aimed to recruit 520 participants. All analyses were performed on an “intention-to-treat (ITT)” method, regardless of whether those in the intervention group read the pamphlet or not. Statistical analyses were computed using *R*, Version 3.5.0^17^

### Masking, randomisation and implementation

Sealed, opaque research envelopes labelled either Group A or B were prepared by two research assistants from the Centre for Eye Research Australia familiar with the conduct of clinical trials. The control pamphlet was folded in half to ensure the research envelopes in both groups were equal in weight and composition. Group allocation of the intervention was determined by the research assistants preparing the envelopes.

A concealed, randomization allocation sequence generation table for two groups, Group A and B, with a 1:1 allocation [2,4,6, block design] was prepared by the statistician (PGS) using *R*, Version 3.5.0^17^ and uploaded to REDCap. Using the REDCap randomization module, upon completion of their baseline survey each participant was automatically randomized to either Group A or B and given their corresponding research envelope by the recruiting researcher (SES). To SES remained masked to group allocation, participants were instructed to not open their research envelope until after leaving the hospital grounds following their antenatal appointment.

All research participants and researchers directly associated with the trial remained masked to participant group allocation until the conclusion of the study and data were analyzed. The study was described to participants as one of maternal and child health, without disclosing the specific focus of the study or the intervention being tested.

### Statistical analysis

Differences among survey responses across treatment groups and follow-up periods were investigated using ordinal logistic mixed model regression analysis. Potentially confounding variables including age, English as a second language at home, previous experience with children, educational attainment, health literacy and past occupational experience in healthcare were adjusted for in the analyses. Specifically, for each outcome measure a ‘Group x Time’ interaction term was assessed and retained in the model if statistically significant, thus indicating a differential response rate across treatment groups at the follow-up compared to baseline visit. Unless otherwise specified, all statistical tests were two-sided, where p ≤0.05 was considered statistically significant.

All participant demographic variables were used to screen for factors that could moderate the outcome of the intervention. Comparison between prima- and multiparous women served as a surrogate for ‘usual care’, providing insights into information parents may have already received or been exposed to in the course of raising their children.

Baseline and follow-up surveys were self-administered using REDCap. This secure, password-protected, web-based data management system is hosted by The University of Melbourne under licence from Vanderbilt University. Each participant’s record was allocated a unique study identifier and data de-identified for statistical analysis.

## RESULTS

### Participant recruitment and study flow

Figure 1 outlines participant recruitment during the study period of 8 March 2018 to 24 June 2018. Of the 692 invited to participate, 518 consented, completed the baseline survey and were randomized to either the intervention or the control group, representing a 74.9% participation rate. The 20 women who ‘failed to proceed’ initially consented but were called to their antenatal appointment prior to completing the baseline survey and did not wish to continue following their appointment. Six participants formally withdrew following randomization and were excluded from the analysis.

**Figure 1:**
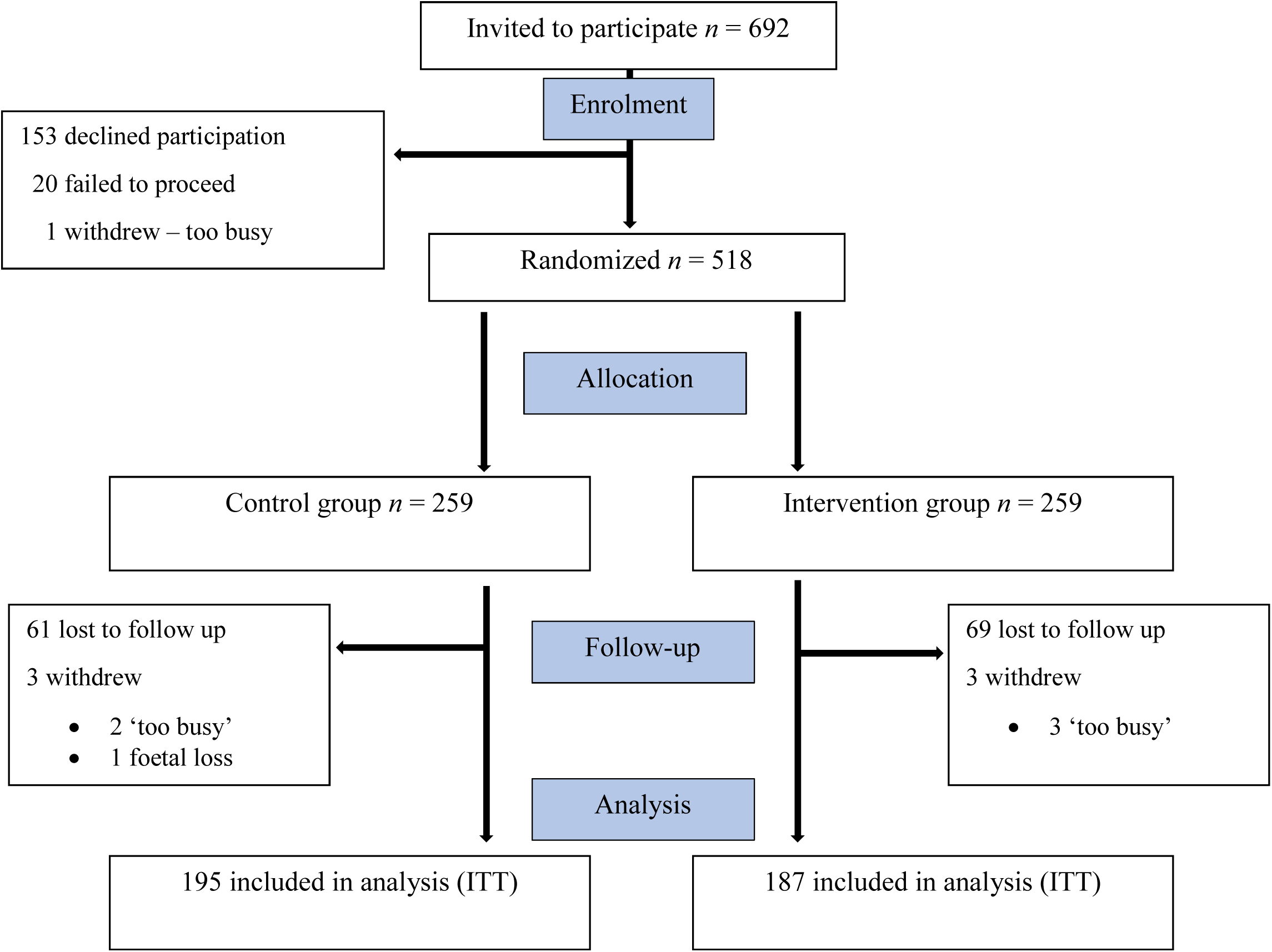
Overview of the study and participant recruitment. Details are displayed as per the CONSORT statement.^28^

Despite receiving reminders as outlined in the protocol, 382/518 women completed the follow-up survey representing a 73.7% overall response rate. Some women attended their antenatal appointment with their partner. When both parents were present, the pregnant woman provided informed consent and completed the survey; however, the partner was neither encouraged nor discouraged from contributing to the decision-making as this could not be controlled for when the participant completed the follow-up survey at home.

### Participant demographics

Generally, participants were tertiary-educated with high health literacy. More than two-thirds of the cohort did not have any other children and few or none had had any previous experience with leukocoria or strabismus. There was no statistically significant difference in demographic characteristics between participants allocated to either the intervention or control group. (Table 1).

**Table 1:**
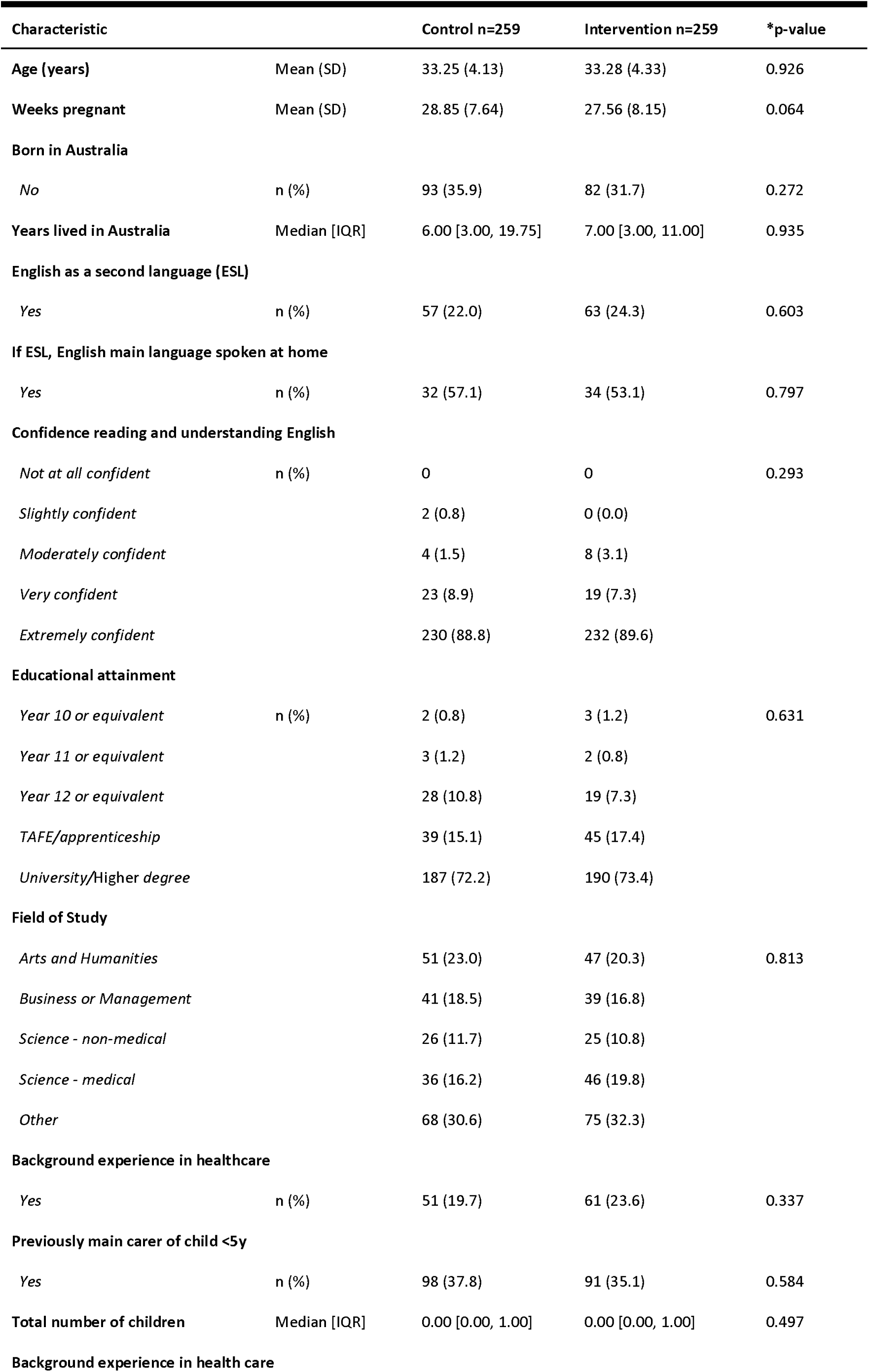
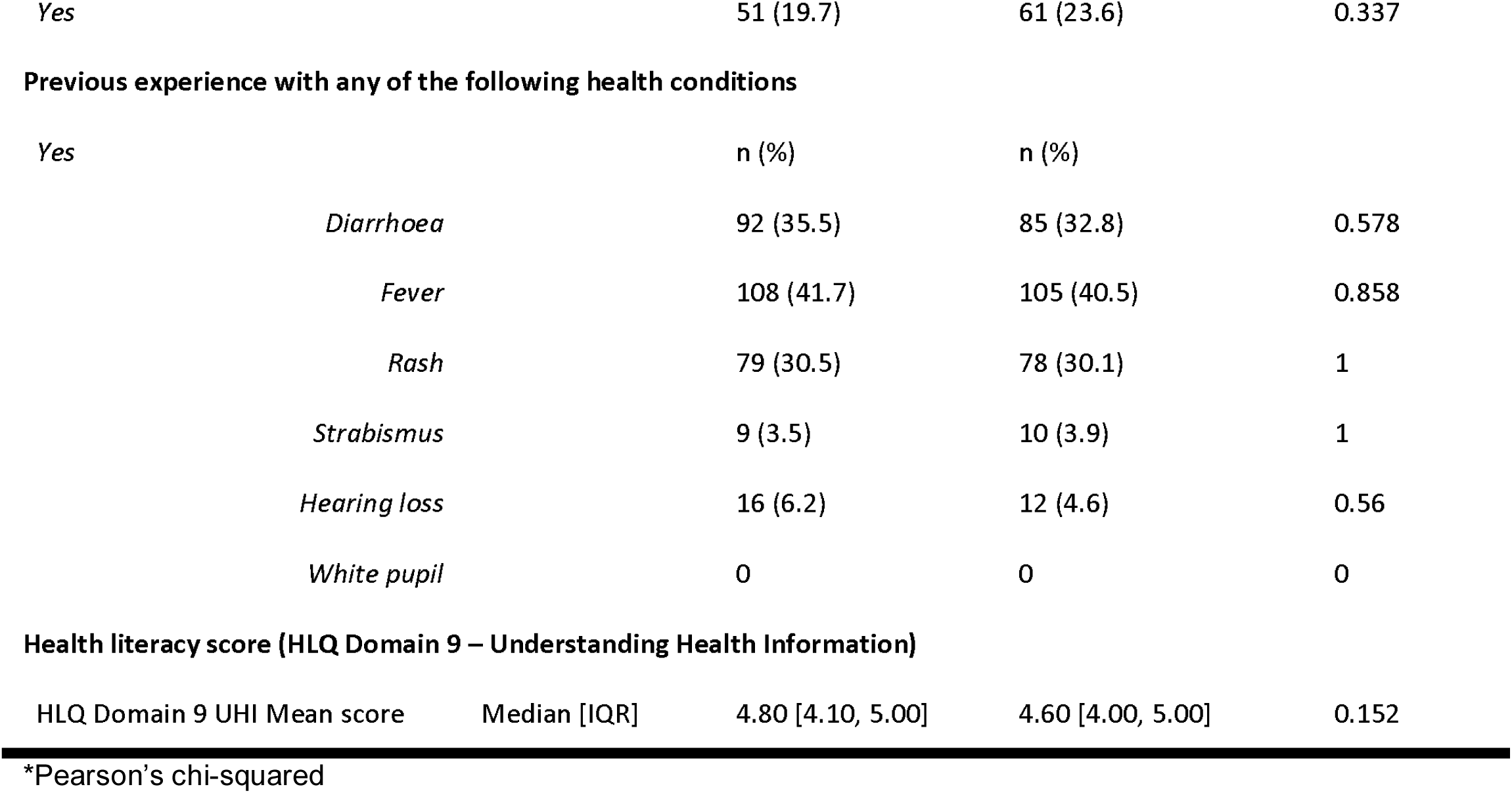
Demographic features of participants allocated to the intervention or control arms.

| Characteristic |  | Control n=259 | Intervention n=259 | *p-value |
| --- | --- | --- | --- | --- |
| **Age (years)** | Mean (SD) | 33.25 (4.13) | 33.28 (4.33) | 0.926 |
| **Weeks pregnant** | Mean (SD) | 28.85 (7.64) | 27.56 (8.15) | 0.064 |
| **Born in Australia** |  |  |  |  |
| *No* | n (%) | 93 (35.9) | 82 (31.7) | 0.272 |
| **Years lived in Australia** | Median [IQR] | 6.00 [3.00, 19.75] | 7.00 [3.00, 11.00] | 0.935 |
| **English as a second language (ESL)** |  |  |  |  |
| *Yes* | n (%) | 57 (22.0) | 63 (24.3) | 0.603 |
| **If ESL, English main language spoken at home** |  |  |  |  |
| *Yes* | n (%) | 32 (57.1) | 34 (53.1) | 0.797 |
| **Confidence reading and understanding English** |  |  |  |  |
| *Not at all confident* | n (%) | 0 | 0 | 0.293 |
| *Slightly confident* |  | 2 (0.8) | 0 (0.0) |  |
| *Moderately confident* |  | 4 (1.5) | 8 (3.1) |  |
| *Very confident* |  | 23 (8.9) | 19 (7.3) |  |
| *Extremely confident* |  | 230 (88.8) | 232 (89.6) |  |
| **Educational attainment** |  |  |  |  |
| *Year 10 or equivalent* | n (%) | 2 (0.8) | 3 (1.2) | 0.631 |
| *Year 11 or equivalent* |  | 3 (1.2) | 2 (0.8) |  |
| *Year 12 or equivalent* |  | 28 (10.8) | 19 (7.3) |  |
| *TAFE/apprenticeship* |  | 39 (15.1) | 45 (17.4) |  |
| *University/Higher degree* |  | 187 (72.2) | 190 (73.4) |  |
| **Field of Study** |  |  |  |  |
| *Arts and Humanities* |  | 51 (23.0) | 47 (20.3) | 0.813 |
| *Business or Management* |  | 41 (18.5) | 39 (16.8) |  |
| *Science - non-medical* |  | 26 (11.7) | 25 (10.8) |  |
| *Science - medical* |  | 36 (16.2) | 46 (19.8) |  |
| *Other* |  | 68 (30.6) | 75 (32.3) |  |
| **Background experience in healthcare** |  |  |  |  |
| *Yes* | n (%) | 51 (19.7) | 61 (23.6) | 0.337 |
| **Previously main carer of child <5y** |  |  |  |  |
| *Yes* | n (%) | 98 (37.8) | 91 (35.1) | 0.584 |
| **Total number of children** | Median [IQR] | 0.00 [0.00, 1.00] | 0.00 [0.00, 1.00] | 0.497 |
| **Background experience in health care** |  |  |  |  |

|  |  |  |  |
| --- | --- | --- | --- |
| Yes | 51 (19.7) | 61 (23.6) | 0.337 |

**Previous experience with any of the following health conditions**
|  |  |  |  |
| --- | --- | --- | --- |
| Yes | n (%) | n (%) |  |
| *Diarrhoea* | 92 (35.5) | 85 (32.8) | 0.578 |
| *Fever* | 108 (41.7) | 105 (40.5) | 0.858 |
| *Rash* | 79 (30.5) | 78 (30.1) | 1 |
| *Strabismus* | 9 (3.5) | 10 (3.9) | 1 |
| *Hearing loss* | 16 (6.2) | 12 (4.6) | 0.56 |
| *White pupil* | 0 | 0 | 0 |
| **Health literacy score (HLQ Domain 9 – Understanding Health Information)** |  |  |  |  |
| HLQ Domain 9 UHI Mean score | Median [IQR] | 4.80 [4.10, 5.00] | 4.60 [4.00, 5.00] | 0.152 |

\*Pearson's chi-squared

### Primary outcomes

Primary outcomes for both leukocoria and strabismus are described in Table 2. At follow-up, participants randomized to the intervention group were more likely to report a higher level of concern when faced with the clinical scenario describing leukocoria in a photograph of their child. (OR 1.711, [CI 1.176,2.497] p=0.005) For all participants, those who searched for more information about leukocoria had an almost 3-fold higher odds of being more concerned (OR 2.778, [CI 1.743,4.455] p<0.001). Participants in the intervention group were less likely to disagree with the statement: *“I would be concerned if I saw a white pupil in a photograph in my baby”* (OR 0.407, [CI 0.267,0.618] p<0.001). Searching for more information about strabismus was associated with a 62% decrease in the odds of disagreeing with this statement (OR 0.379, [CI 0.211,0.672] p=0.001) Moreover, participants referring to their pamphlet when completing the follow-up survey were less likely to disagree. (OR 0.541, [CI 0.309,0.941] p=0.030)

**Table 2:** Level of concern if leukocoria or strabismus observed

| Variable | OR | 95% CI<br>[LCI,UCI] | *p-value |
| --- | --- | --- | --- |
| **Clinical scenario: Leukocoria - Level of concern** |  |  |  |
| Age | 1.017 | [0.972,1.063] | 0.470 |
| Intervention group | 1.711 | [1.176,2.497] | 0.005 |
| Searched for more<br>information-white pupil | 2.778 | [1.743,4.455] | <0.001 |
| **Statement of concern: Leukocoria**<br>**“I would be concerned if I saw a white pupil in a photograph of my baby”** |  |  |  |
| Age | 0.980 | [0.034,1.028] | 0.400 |
| Intervention group | 0.407 | [0.267,0.618] | <0.001 |
| Searched for more<br>information-strabismus | 0.379 | [0.211,0.672] | 0.001 |
| Referred to pamphlet for<br>follow-up survey | 0.541 | [0.309,0.941] | 0.030 |
| **Clinical scenario: Strabismus - Level of concern** |  |  |  |
| Age | 0.992 | [0.945,1.042] | 0.749 |
| Intervention group | 1.310 | [0.886,1.940] | 0.176 |
| Year level of educational attainment |  |  |  |
| Year 9 | 0.992 | [0.207,4.336] | 0.992 |
| Year 10 | 1.045 | [0.499,2.149] | 0.905 |
| Year 11 | 2.294 | [1.336,3.950] | 0.003 |
| Searched for more<br>information-strabismus | 2.009 | [1.128,3.579] | 0.018 |
| **Statement of concern: Strabismus**<br>**“I would be concerned if I saw my 2-year-old baby’s eye turning (cross-eyed) some of the time”** |  |  |  |
| Age | 1.038 | [0.985,1.093] | 0.161 |
| Intervention group | 0.747 | [0.482,1.156] | 0.191 |
| Referred to pamphlet for<br>follow-up survey | 0.400 | [0.225,0.707] | 0.002 |

\*Ordinal mixed model regression OR=Odds Ratio CI = confidence interval

For the clinical scenario describing strabismus in a child, there was no statistically significant difference in the level of concern between the intervention and control groups at follow-up. However, participants who looked up more information about strabismus at follow-up had a 2-fold increase in the odds of being more concerned about this sign. (OR 2.009, [CI 1.128,3.579] p=0.018). Moreover, participants referring to their pamphlet to complete the follow-up survey were less likely to disagree with the statement: *“I would be concerned if I saw my 2-year-old baby’s eye turning (cross-eyed) some of the time”* (OR 0.400, [CI 0.225,0.707] p=0.002).

### Secondary outcomes

As shown in Table 3, responding to the clinical scenario for leukocoria, participants in the intervention group were less likely to delay seeking health advice (OR 0.560, [CI 0.382,0.817] p=0.003), as were participants who searched for more information on strabismus (OR 0.427, [CI 0.248,0.725] p=0.002). Responding to the clinical scenario for strabismus, for all participants there was a 2.7-fold increased odds of seeking advice sooner at follow-up (OR 2.744, [CI 1.384,5.442] p=0.004). Whilst group allocation in isolation did not have a predictive effect on help-seeking intention for the strabismus scenario, an interaction effect between follow-up visit and group allocation was observed, whereby at follow-up, participants who received the intervention were less likely to delay seeking advice (OR 0.318, [CI 0.125,0.806] p=0.016). Moreover, participants who spoke a language other than English at home were more likely to seek advice promptly (OR 0.218 [CI0.098,0.482] p<0.001).

**Table 3:** Help-seeking intention if leukocoria or strabismus are observed.

| Variable | OR | 95% CI<br>[LCI, UCI] | *p-value |
| --- | --- | --- | --- |
| **Help-seeking Intention - Leukocoria** |  |  |  |
| Age | 0.982 | [0.939,1.028] | 0.436 |
| Intervention group | 0.560 | [0.382,0.817] | 0.003 |
| Searched for more information-strabismus | 0.427 | [0.248,0.725] | 0.002 |
| **Help-seeking Intention - Strabismus** |  |  |  |
| Age | 1.001 | [0.925,1.082] | 0.986 |
| Follow-up survey | 2.744 | [1.384,5.442] | 0.004 |
| Intervention group | 1.341 | [0.643,2.796] | 0.433 |
| Language other than English<br>primarily spoken at home | 0.218 | [0.098,0.482] | <0.001 |
| Follow-up survey: intervention<br>group | 0.318 | [0.125,0.806] | 0.016 |

*\*Ordinal mixed model regression OR= Odds Ratio*

## DISCUSSION

This study demonstrated that providing women with appropriate information about leukocoria and strabismus could improve their knowledge and guide their help-seeking intentions. Specifically, participants in the intervention group were more likely to demonstrate an increased level of concern and were more likely to seek advice quickly if they observed leukocoria. However, if they observed strabismus, the intervention group’s level of concern did not differ, although their intended time to help-seeking was significantly shorter than the control group. Neither age of the participants nor whether they had other children predicted their level of concern or help-seeking intention. Previous studies has revealed that providing health information to parents can lead to improved health outcomes in their children.^18–21^

Compared to the control group, those in the intervention group had a significant increase in their level of concern and help-seeking intention if they observed leukocoria, demonstrating the positive effect of the intervention for this sign. Given group allocation did not predict a participant’s level of concern for strabismus likely suggests they were already familiar with this condition. However, the highly significant interaction effect observed between the intervention group at follow-up positively predicted help-seeking intention for this sign. Here, we might conclude that whilst participants were already aware that strabismus could occur in children, they were unfamiliar with the urgency with which medical advice should be sought and the intervention appropriately guided their help-seeking behaviour.

Few participants confirmed searching for more information after completing their baseline survey and it is possible participants under-reported their self-directed research, which could bias these results. Curiously, self-directed research on white pupils as well as strabismus predicted an increase in their level of concern if they observed leukocoria. People who observe photo-leukocoria may regard it as an artefact caused by a camera flash or room lighting and thus overlook its potential seriousness.^22^ Therefore, in this RCT, either some participants also did not think leukocoria could be a significant finding, remaining unmotivated to search for information, or the information encountered about leukocoria was not medically informative. It is not clear why searching for strabismus might have predicted an increased level of concern for leukocoria. We postulate that information encountered during their searches about strabismus may have referenced leukocoria, and that when the two occur together, prompt attention is imperative.

Whilst few participants reported referring to their pamphlet or the internet to answer their follow-up survey, it is notable that participants in the intervention group were more likely to refer to their pamphlet. This could suggest they recalled the information was contained in their pamphlet, unlike the control group who referred to the internet. Moreover, referring to their pamphlet was weakly associated with increasing levels of concern for leukocoria and strabismus as well as predicting help-seeking intention for leukocoria. It is possible that the pamphlet clarified for the reader when strabismus might be normal or pathological, giving rise to poor vision or a sign of more sinister disease. Despite these findings, the numbers were small, and it is possible not all participants reported referring to their pamphlet to complete the follow-up survey.

An equal number of participants in both the control and intervention groups already had children. One might expect that those who already had children would have received any currently provided pediatric eye-health information; may have been exposed to information via print or social media; or that they might have been motivated to seek out information themselves if they felt it was required. Given a participant’s previous experience with children did not predict level of concern or help-seeking behaviour confirms that information about leukocoria or strabismus is not currently provided.

The observed changes in the level of concern and help-seeking intention may be attributed to the Question-Behaviour Effect, whereby merely being asked a question, individuals will over predict their behaviour.^23^ That is, faced with a hypothetical scenario, the individual would seek help sooner than if faced with the same situation in real life. Studies reporting delayed diagnosis of retinoblastoma attribute parents’ slow response to strabismus because it is familiar or they expect it to resolve spontaneously; and leukocoria because it is not obvious or visible all the time.^24^ Together with the fact the child is otherwise well and is unlikely to demonstrate an overt sign of poor vision, parents delay seeking advice. Thus, one cannot be certain that the hypothetical responses observed in the RCT reported herein would necessarily translate to parents actually seeking health advice promptly.

### Limitations

The single-centre study design may have unintentionally biased the outcomes. Although participants’ demographic characteristics were equally distributed between the two groups and were adjusted for confounding variables, their homogeneity may have contributed to the positive outcomes observed in this study. Strong literacy skills and command of the English language may have enabled them to engage more fully with, and understand the contents of, the intervention.

In this study, fathers or partners contributing to the RCT could neither be prevented nor controlled. Thus, it is not known whether a difference of opinion may have tempered or intensified their concern or help-seeking intention. Other than for mental health and behaviour^25^, few studies have explored a father’s (or partner’s) role in child health and help-seeking compared to the mother’s.^26^ Although no significant differences between mothers’ and fathers’ knowledge and attitudes towards seeking eye-health care for their children have been identified^27^, the distinction between maternal and paternal help-seeking behaviour is not usually made, referring only to ‘parental’ behaviour.^28–30^ Future studies could consider comparing responses between both parents to determine whether there are any differences, as these would need to be addressed to optimize health promotion initiatives.

The time between receipt of their pamphlet and the follow-up survey was short (2 weeks). Thus, participants may have had a better recall or retention of information than would normally occur. Nonetheless, this study did establish that providing information that can be understood and interpreted by parents can improve knowledge and help-seeking behaviour. This study could be extended in the future by repeating the follow-up survey at a later time-point to better understand parent recall of information and effect on help-seeking intention.

## CONCLUSION

Although these study findings are not generalizable, they do provide significant, previously unreported insights into community knowledge of leukocoria and strabismus, and help-seeking intentions if observed. For pediatric eye diseases with presenting signs of leukocoria or strabismus, educating parents to remain vigilant and how to respond may result in earlier diagnosis, treatment and improved outcomes.

## Data Availability

All data available upon request

## Abbreviations

ANZCTR: Australia and New Zealand Clinical Trials Registry
CI: Confidence interval
HLQ: Health Literacy Questionnaire
IMB: Information-Motivation-Behavioural Skills
NHMRC: National Health and Medical Research Council
OR: Odds ratio
RCT: Randomized Controlled Trial
REDCap: Research Electronic Data Capture

## ACKNOWLEDGMENTS

We thank all the women who agreed to participate in this study and those parents who assisted with the development of the intervention. Further, we acknowledge Carly Parfett, Tanya Pejnovic, Ms Lisa Kearns and Ms Linda Clarke for administrative assistance, and Ms Lori Bonertz for editorial assistance in the preparation of this manuscript. We also thank MCATS (Melbourne Clinical and Translational Sciences research platform) for the administrative and technical support that greatly facilitated this research.

---

Checklist of items to include when reporting a randomized trial (56-58)
| PAPER SECTION<br>And topic | Item | Description | Reported<br>on page # |
| --- | --- | --- | --- |
| *TITLE & ABSTRACT* | 1 | How participants were allocated to interventions (e.g., "random allocation", "randomized", or "randomly assigned"). | 1,4 |
| *INTRODUCTION*<br>Background | 2 | Scientific background and explanation of rationale. | 5 |
| *METHODS*<br>Participants | 3 | Eligibility criteria for participants and the settings and locations where the data were collected. | 6 |
| Interventions | 4 | Precise details of the interventions intended for each group and how and when they were actually administered. | 7 |
| Objectives | 5 | Specific objectives and hypotheses. | 6, 9 |
| Outcomes | 6 | Clearly defined primary and secondary outcome measures and, when applicable, any methods used to enhance the quality of measurements (e.g., multiple observations, training of assessors). | 9 |
| Sample size | 7 | How sample size was determined and, when applicable, explanation of any interim analyses and stopping rules. | 9 |
| Randomization --<br>Sequence generation | 8 | Method used to generate the random allocation sequence, including details of any restriction (e.g., blocking, stratification). | 10 |
| Randomization --<br>Allocation concealment | 9 | Method used to implement the random allocation sequence (e.g., numbered containers or central telephone), clarifying whether the sequence was concealed until interventions were assigned. | 10 |
| Randomization --<br>Implementation | 10 | Who generated the allocation sequence, who enrolled participants, and who assigned participants to their groups. | 9, 10 |
| Blinding (masking) | 11 | Whether or not participants, those administering the interventions, and those assessing the outcomes were blinded to group assignment. When relevant, how the success of blinding was evaluated. | 9, 10 |
| Statistical methods | 12 | Statistical methods used to compare groups for primary outcome(s); Methods for additional analyses. such as subgroup analyses and adjusted analyses. | 10, 11 |
| RESULTS<br>Participant flow | 13 | Flow of participants through each stage (a diagram is strongly recommended). Specifically, for each group report the numbers of participants randomly assigned, receiving intended treatment, completing the study protocol, and analyzed for the primary outcome. Describe protocol deviations from study as planned, together with reasons. | Figure 1 |
| Recruitment | 14 | Dates defining the periods of recruitment and follow-up. | 11 |
| Baseline data | 15 | Baseline demographic and clinical characteristics of each group. | Table 1 |
| Numbers analyzed | 16 | Number of participants (denominator) in each group included in each analysis and whether the analysis was by "intention-to-treat". State the results in absolute numbers when feasible (e.g., 10/20, not 50%). | 11 |
| Outcomes and estimation | 17 | For each primary and secondary outcome, a summary of results for each group, and the estimated effect size and its precision (e.g., 95% confidence interval). | Table2&3 |
| Ancillary analyses | 18 | Address multiplicity by reporting any other analyses performed, including subgroup analyses and adjusted analyses, indicating those pre-specified and those exploratory. | n/a |
| Adverse events | 19 | All important adverse events or side effects in each intervention group. | n/a |
| DISCUSSION<br>Interpretation | 20 | Interpretation of the results, taking into account study hypotheses, sources of potential bias or imprecision and the dangers associated with multiplicity of analyses and outcomes. | 14-16 |
| Generalizability | 21 | Generalizability (external validity) of the trial findings. | 17-18 |
| Overall evidence | 22 | General interpretation of the results in the context of current evidence. | 18 |

